# Proteomic deconvolution reveals distinct immune cell fractions in different body sites in SARS-Cov-2 positive individuals

**DOI:** 10.1101/2022.01.21.22269631

**Authors:** Javan Okendo, David Okanda, Peter Mwangi, Martin Nyaga

**Author notes:** Corresponding author Javan Ochieng Okendo, Systems and Chemical Biology Division, Department of Integrative Biomedical Sciences, Institute of Infectious Disease and Molecular Medicine, Faculty of Health Sciences, University of Cape Town, Anzio Road Observatory, Cape Town, 7925, South Africa.

## Abstract

**Background:** Severe acute respiratory syndrome coronavirus 2 (SARS-CoV-2) continues to be a significant public health challenge globally. SARS-CoV-2 is a novel virus, and what constitutes immunological responses in different human body sites in infected individuals is yet to be presented. We set to determine the various immune cell fractions in gargle solution, bronchoalveolar lavage fluid, nasopharyngeal, and urine samples post-SARS-CoV-2 infection in humans.

**Materials and methods:** We downloaded proteomics data from (https://www.ebi.ac.uk/pride/) with the following identifiers: PXD019423, n=3 (gargle solution), PXD018970, n=15 (urine), PXD022085, n=5 (Bronchoalveolar lavage fluid), PXD022889, n=18 (nasopharyngeal). MaxQuant was used for the peptide spectral matching using humans, and SARS-CoV-2 was downloaded from the UniProt database (Access date 9^th^ January 2022). The protein count matrix was extracted from the proteins group file and used as an input for the cibersort for the immune cells fraction determination.

**Results:** The body of individuals infected with the SARS-CoV-2 virus is characterized by different fractions of immune cells in Bronchoalveolar lavage fluid (BALF), nasopharyngeal, urine, and gargle solution. BALF has more abundant memory B cells, CD8, activated mast cells, and resting macrophages than urine, nasopharyngeal, and gargle solution. Our analysis also demonstrates that each body site comprises different immune cell fractions post-SARS-CoV-2 infection in humans.

**Conclusion:** Different body sites are characterized by different immune cells fractions in SARS-CoV-2 infected individuals. The findings in this study can inform public health policies and health professionals on treatment strategies and drive SARS-CoV-2 diagnosis procedures.

## 1 Introduction

Acute respiratory syndrome coronavirus 2 (SARS-CoV-2) infection continues to cause significant global public health due to the constant emergence of new variants, which makes it had to control (1). Currently, there is no approved antiviral drug to treat SARS-Cov-2 infection, and the available vaccines do not have 100% protection against the presently circulating variants. COVID-19 may result in clinical features such as cardiovascular, neurological, thrombosis, and renal failure (2). Approximately 298 million COVID-19 cases with over 5.4 million deaths were reported globally on 08-01-2022 (https://CoVid19.who.int/). Globally, the exact impact of SARS-CoV-2 infection is unknown even though the number of confirmed cases is still in the upward trajectory. The number of cases seems to vary geographically. As of 09 January 2022, the approximate number of cases per region were as follows America: 108 million, Europe: 108 million, Southeast Asia: 45 million, Eastern Mediterranean: 17 million, Western pacific: 11 million, and Africa: 7.5 million (https://CoVid19.who.int/). SARS-CoV-2 is mainly the disease of the lungs, but it also targets the small vessels, immune organs such as lymph nodes, resulting in systemic vasculitis, and perturbed immune functions. SARS-CoV-2 infection in the lungs causes alveolar damage-causing hyaline membrane formation around the affected alveolar (3). The alveolar damage is approximated to cause ∼10% in individuals infected with the SARS-CoV-2 virus globally (4).

COVID-19 spread from person to person through direct contact or encountering infected surfaces. When SARS-CoV-2 is inhaled, it enters the human host cells via angiotensin-converting enzyme 2 (ACE2) receptors (5). Once the virus enters the human cells, it starts replicating, leading to population expansion (5). While in the cells, it induces the local immune cells to begin producing cytokines and chemokines, resulting in the attraction of other immune cells in the lung and the other infected body sites, which causes excessive tissue damage (6). A growing body of evidence indicates that the SARS-CoV-2 virus is not confined in the human lungs. Still, it also affects the other body organs, such as the kidney, where it causes acute kidney injury (AKI) (7). In other individuals infected with SARS-CoV-2, neurological, cardiovascular, and intestinal malfunctions have also been reported (2).

Understanding what constitutes immune response in SARS-CoV-2 infected individuals, studies have been carried out to understand the different cell populations from various body sites. Koch et al. 2021, used RNAseq deconvolution to understand the immune response in children and adults infected with the SARS-CoV-2 virus (8). Grant et al. 2021 used bulk RNAseq deconvolution to study the cell populations using BALF as their biospecimen in SARS-CoV-2 infected individuals. They concluded that BALF is enriched in T cells and memory cells in individuals infected with SARS-CoV-2 (9). The body elicits a unique immune response following SARS-CoV-2 infection. This was revealed using the deconvolution of the RNAseq data by Butler et al. 2021, where they showed enrichment of cells spanning goblet, ciliated airway, and epithelial cells (10). A combination of single-cell and bulk RNA profiling has also been used to reveal that the alveolar space of individuals infected with SARS-CoV-2 is more enriched with macrophages and T cells without neutrophils which are the primary defense cells in the lungs (11). Using proteomic deconvolution, Voss et al. 2021 demonstrated that neutralizing antibodies responses are largely directed against the epitopes such as S protein in SARS-CoV-2 infected individuals (12).

The studies highlighted above have used transcriptomic data to understand the diverse cell immune populations implicated in response to SARS-CoV-2 infection in humans. There is a negative correlation between proteomics and transcriptomics. What is expressed may not be translated to proteins, the molecule responsible for conduction the organism’s actual biological function. The only proteomic deconvolution study mentioned above was used to investigate the neutralizing antibodies against SARS-CoV-2. To the best of our understanding, none of the studies has been conducted to understand the different immune cell populations using proteomic data extracted from nasopharyngeal, bronchoalveolar lavage fluids, gargle solution, and urine samples. On this basis, we set to carry out proteomic deconvolution of these data obtained from different human sites to understand the immunological dynamics in individuals infected with the SARS-CoV-2 virus. The findings in this study can inform public health and health professionals on treatment strategies and drive SARS-CoV-2 diagnosis procedures.

## 2 Materials and methods

This study analyzed publicly available data downloaded from the Protein Identification Database, PRIDE (https://www.ebi.ac.uk/pride/) repository. The data used in this analysis had the following identifiers: PXD019423 (gargle solution, n=3) (13), PXD018970 (Urine, n=15) (14), PXD022085 (BALF, n=5) (15), and PXD022889 (nasopharynx, n=18) (16). We acknowledge that the samples used in this study were processed in different laboratories worldwide, and the sample preparation protocols have been reported elsewhere in the respective publications (13,15–17).

### 2.1 Bioinformatics analysis of raw tandem (MS/MS) mass spectrometry proteomic data

Raw data files were processed with MaxQuant version 1.6.10.43 (18) for protein and peptide identification using the Andromeda search engine and the combined Uniprot proteome for *Homo sapiens* (Proteome ID: UP000005640, 78120 entries, and SARS-CoV-2 Proteome ID: UP000464024, entries 17 both accessed on 08/01/2022). MaxQuant default parameter settings were used for the MS/MS database search, with carbamidomethylation of cysteine residues and acetylation of protein N-termini selected as fixed modification and oxidation of methionine as variable modification. The peptide spectral matches (PSMs) were filtered at a 1 % false discovery rate (FDR), and the precursor mass tolerance was set at 20 ppm. Trypsin/P was selected as protease, label-free quantitation (LFQ) was enabled. The samples from the different studies were processed together to ensure cross normalization, making the proteome comparison and deconvolution more accurate. Reverse hits and common contaminants were removed using Bioconductor package ‘Differential Enrichment analysis of Proteomics data’ version 1.2.0 (19) before the downstream analysis. The protein groups identified in 70% of patients by at least two unique peptides were retained for analysis. Post-processing of the data, we kept 2443 proteins that satisfied our data quality control threshold, which was further used in the downstream analysis and deconvolution.

### 2.2 Proteomic deconvolution

Cibersort (20) was used to do the immune cell deconvolution. Briefly, a signature matrix consisting of 22 immune cells was used as a reference matrix. The protein mixture matrix obtained from MaxQuant (18) was used as an input. Batch correction mode (B-mode) was selected. The pipeline was run on the absolute mode to scale the relative cellular fractions into the score reflecting each cell type absolute proportion in a mixture. Finally, the permutation for significance analysis was set to 1000. Cell expression imputation was conducted using the following parameters: the LM22 was used as a signature matrix and the proteomic count matrix as a mixture counts data frame. The batch correction mode (B-mode) was activated. ComplexHeatmaps package was used to generate heatmaps and the “prcomp” function with doing the principal component analysis (PCA) in R version 4.1.

## 3 Results

The previous research has demonstrated that the angiotensin-converting enzyme 2 (ACE2) is the entry point of SARS-CoV-2 into human cells (4,21). SARS-CoV-2 is selective because it only infects the cell expressing the ACE2 proteins, and it loses the ability to infect the non-ACE2 expressing cells, as was demonstrated by Zhou et al. 2020 (22). This study hypothesized that different body sites would be characterized by different immune responses due to the differential ACE2 protein expression in various body sites. Thus, we conducted proteomic deconvolution to determine cell population in proteomic data obtained from other human body sites following SARS-CoV-2 infections.

### 3.1 Unique sets of immune cells characterize each human body site following SARS-CoV-2 infection

There have been several attempts to understand what constitutes immune response post-SARS-CoV-2 infection in humans, but a universal understanding of immune dynamics is yet to be presented. Our analysis shows some similarity between immune cells available in the BALF and urine samples due to the overlap observed in the PCA [Figure 1]. Interestingly, the nasopharyngeal immune cell fraction is very different from the BALF and nasopharyngeal samples [Figure 1]. The immune cells detected in the gargle solution were also seen in BALF, urine, and nasopharyngeal proteomic samples, as shown in [Figure 1]. A plausible reason why the gargle solution does not have a specific clustering pattern is daily temporal dynamics in the buccal cavity micro and macroenvironment.

**Figure 1:**
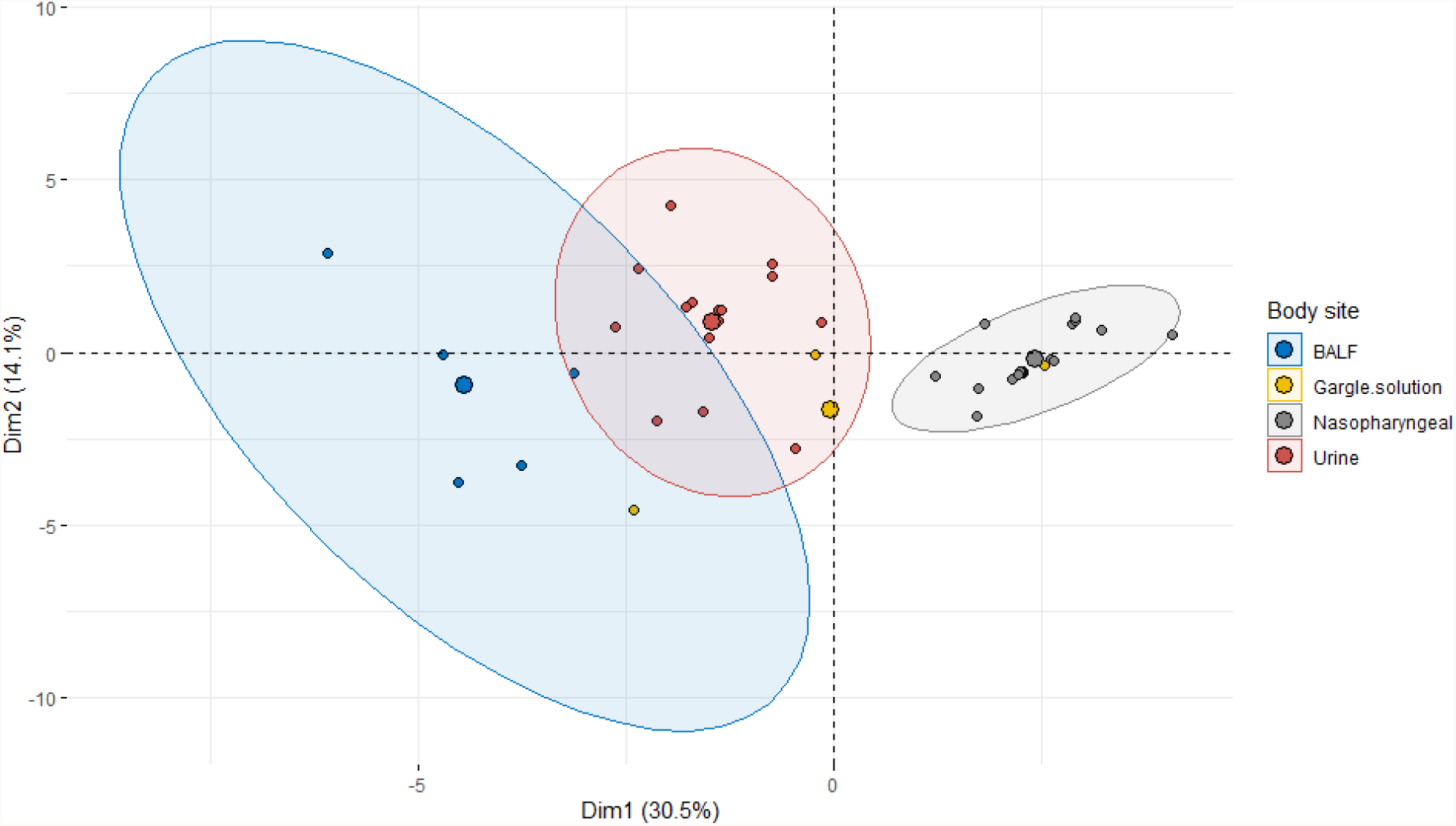
Principal component analysis showing clustering of immune cells identified from different body sites from individuals infected with SAR-CoV-2, BALF (bronchoalveolar lavage fluid).

### 3.2 Different proportions of immune cells characterize bronchoalveolar lavage fluid, gargle solution, nasopharyngeal and urine samples

In Figure *2*, we assessed the immune cell profiles in individuals infected with SARS-CoV-2. Individuals demonstrated heterogeneity in the proportion of immune cells and indication that robustness of immune response is not the same in SARS-CoV-2 patients. BALF samples had a high proportion of activated mast cells, alveolar macrophages (M0), CD8 T cells, and memory B cells [Figure *2*]. Gargle solution samples had a high proportion of naïve CD4 cells, regulatory T cells, memory B cells, and activated natural killer cells [Figure *2*]. A good number of nasopharyngeal samples had a high proportion of CD8 T cells, memory B cells, resting CD4 cells, activated mast cells, monocytes, and plasma cells [Figure *2*]. In urine samples, the proinflammatory macrophages M1 were present in most of the individuals [Figure *2*]. The activated mast cells were also present in the urine samples at a higher proportion, together with gamma and delta T cells [Figure *2*]. Interestingly, the proportions of the memory B cells were lower than the nasopharyngeal, gargle solution, and BALF proteomic samples we compared in our analysis [Figure *2*]. The follicular helper T cells were also detected in several urine samples.

**Figure 2:**
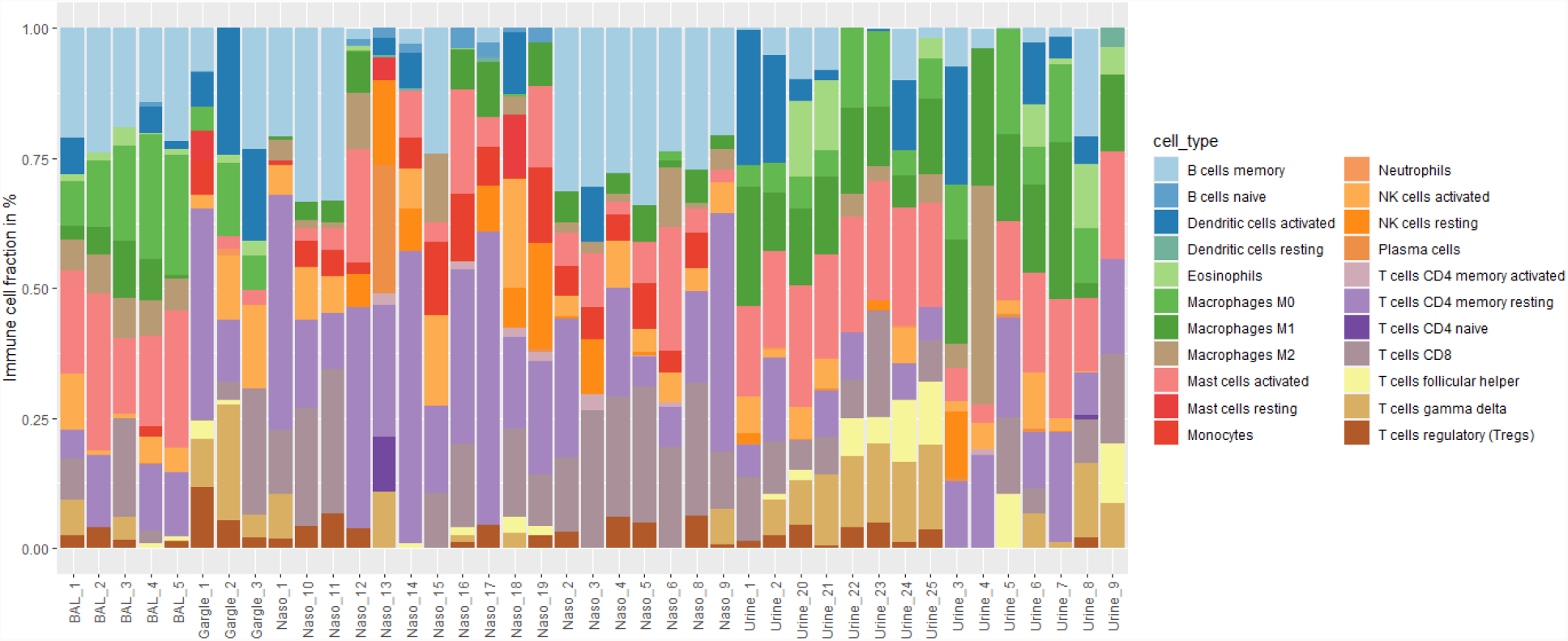
Stacked barplot showing the proportions of immune cells identified from proteomic samples obtained from gargle solution, nasopharyngeal, BALF, and urine samples

### 3.3 The cluster of Differential 8 (CD8) and memory B cells are the main cells involved in controlling SARS-CoV-2 replication in human lungs

The relative immune cell abundance was investigated in our samples. We interestingly showed that the main immune response is more pronounced in the alveolar space in the human lungs [Figure 3]. Our analysis demonstrated two main clusters of the immune cells involved in response to SAR-CoV-1 infection in humans [Figure 3]. Cluster 2 comprises macrophage M0 (resting), activated mast cells, CD8 T cells, and memory B cells. These cells (in cluster 2) are more abundant mostly in the bronchoalveolar lavage fluid samples but less abundant in the urine, gargle solution, and nasopharyngeal samples [Figure 3]. In our data, we observed overlap in the cell population in different proteomic data we analyzed. The same cells in the lungs are also detected in the urine and gargle solution. We also observed clustering of one gargle solution sample with a cell composition like the one detected in the nasopharyngeal samples in our data [Figure 3]. There were three main clusters with small overlaps in the samples in the data because most of the samples from a given body site clustered closely together in each dendrogram in our data [Figure 3]. The first clustered comprised both cellular and innate immune cells, and their abundance was low in all the samples we analyzed. Some proteins immune cells identified in the bronchoalveolar lavage fluids (BALF) were also present in the urine samples, though in less abundance than the proteomic data obtained in the alveolar space.

**Figure 3:**
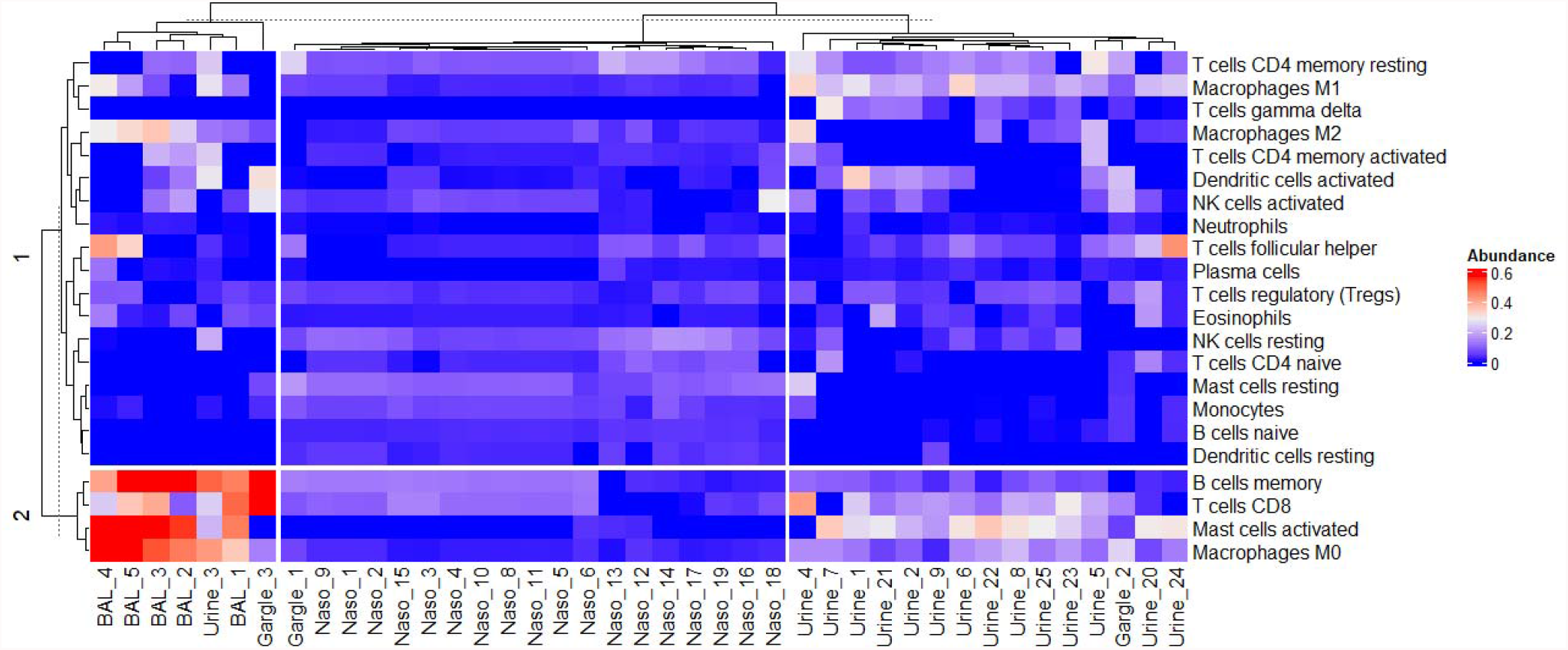
Heatmap showing the relative abundance of immune cells in proteomic samples collected from different human body sites.

## 4 Discussion

MultiOmics approaches have been employed to understand the pathogenesis and immune response following SARS-CoV-2 infections in humans (23–25). These studies have been conducted to understand the main biomarkers and potential drug targets to control the spread of novel SARS-CoV-2 virus globally. Our previous work (25) demonstrated that SARS-CoV-2 infection results in a considerable difference in proteome diversity in different body sites. The clinical manifestations of SARS-CoV-2 infection include fever, headache, cough, muscle pain, diarrhea, and myocarditis, among others (21). Previous studies have demonstrated that the muscle pains that characterize SARS-CoV-2 infection in humans are caused by the cytokine storm (26). The primary source of cytokine in the infected individuals is the infected macrophages and the lung epithelial cells (26). We observed differential immune response in different body sites, as demonstrated by the immune cells’ abundance in gargle solution, nasopharyngeal, urine, and bronchoalveolar lavage fluid. And this finding was consistent with the Hamming et al. 2004 findings. They demonstrated that the angiotensin-converting enzyme 2 (ACE2) is expressed differently in different bodies, hence the differential immune response following SARS-CoV-2 infections in humans (4). Our data shows that the memory B cells, Cluster of Differential 8 (CD8) cells, activated mast cells, and the resident macrophages are more abundant in the BALF than in the urine, gargle solution, and nasopharyngeal samples [Figure *3*]. And a plausible explanation for this is due to the high expression of ACE2 proteins in the lung epithelial compared with the other body sites we compared. Study demonstrate that the immunological cells involved in response to SARS-CoV-2 are mentioned above. Our data also demonstrates that the immune cells that characterize different body sites are unique, and each body site responds differently to the virus once it enters the body. Our study demonstrates that the urine samples have interesting immune cell fractions involved in the innate and cellular immune responses. Our data indicate that the critical immune response at the site of infection is primarily cellular because it is only the cellular immune cells that are more abundant in the BALF samples. On the other hand, the individuals’ immune cells profiles indicated that the urine samples contain essential immune cells, warrants further investigation. We opine that these proteins detected in the urine have theory source from the kidney, an organ that is affected by SARS-CoV-2 in critically ill patients (7,27). In general, our analysis demonstrated that the CD4 T cell and Macrophage immune responses in individuals with SARS-CoV-2 infection is impaired. This finding is in agreement with the Evangelos et al 2020 (28) finding where they demonstrated that activated memory B cells are upregulated in SARS-CoV-2 infected individuals compared with the control samples (28).

The studies conducted around gargle solutions have demonstrated the presence of SARS-CoV-2 related peptides (13). In [Figure *1*], our data show that the gargle solutions immune cells do not have a specific proteome profile because they do not have a particular cluster. The lack of specific clustering can be attributed to the different macro and microenvironment conditions in the buccal cavity. Interestingly, our data show some similarity in the cellular population in the urine samples and the BALF fluid. This overlap could be due to the drainage from the kidneys, where it has been demonstrated that SARS-CoV-2 causes acute kidney injury (AKI) (7) hence the similarity observed in Figure 1 and Figure *3*Figure *3*.

In conclusion, our data demonstrated that the immune response in individuals infected with SARS-CoV-2 is not the same. This is a proof-of-concept study, demonstrating that SARS-CoV-2 therapeutics and drugs should be designed to target specific cells, specifically lung epithelial cells. These are the cells targeted by the SARS-CoV-2 virus due to their overexpression of ACE2 proteins. We showed that human immune response post-SARS-CoV-2 infection is differential and not the same in different body sites.

Study limitations include different sample preparation protocols, which can be a source of a possible confounder.

## Data Availability

The proteins group file is available on Fig share with DOI 10.6084/m9.figshare.18264890.

https://doi.org/10.6084/m9.figshare.18264890.v1

## 4.1 Competing interest

The authors declare that they have no competing interests.

## 4.2 Consent for publication

Not applicable.

## 4.3 Ethics approval and consent to participate

Not applicable.

## 4.4 Funding

This research did not receive any specific grant from funding agencies in the public, commercial, or not-for-profit sectors.

## 4.5 Authors’ contribution

JO analyzed the proteomic data and wrote the manuscript. DO, MN, PM, and HO wrote the manuscript. All authors reviewed and approved the final draft of the manuscript.

## 4.6 Acknowledgment

We acknowledge all the participating health facilities where the respective data were collected. We also acknowledge the authors of the primary data we used for my reanalysis. Computations were performed using facilities provided by the University of Cape Town’s ICTS High-Performance Computing team: *<u>hpc</u>*.*<u>uct</u>*.*<u>ac</u>*.*<u>za</u>*.

## 4.7 Availability of data and materials

The proteomic data sets used in this study are available on PRIDE with the following accession numbers: PXD019423, PXD018970, PXD022085, and PXD022889. The proteins group file is available on Fig share with DIO 10.6084/m9.figshare.18264890.

## Notes

### Competing Interest Statement

The authors have declared no competing interest.

### Funding Statement

This study did not receive any funding

